# Evaluation of Intact Fish Skin Grafts Plus Standard Care in the Treatment of Venous Leg Ulcers: an interim analysis of the THOR trial

**DOI:** 10.1101/2025.10.29.25339062

**Authors:** Thomas Serena, Brianna Tramelli, Barkley Booker, Emily King, Dereck Shi, Zwelithini Tunyiswa, John C. Lantis

## Abstract

**Background:** Venous leg ulcers (VLUs) are nonhealing wounds that pose considerable clinical and economic challenges. Healing outcomes with existing standard-of-care (SOC) remain limited, creating a pressing need for more effective therapeutic options

**Methods:** An interim analysis of this randomized controlled multicenter clinical trial evaluated intact fish skin graft (IFSG) and SOC versus SOC alone in nonhealing VLUs. The primary endpoint was the percentage of target ulcers achieving complete wound closure in 12 weeks, defined as 100% re-epithelialization without drainage for two consecutive weeks, confirmed by blinded independent review.

**Results:** The statistical analysis revealed that the treatment arm improved full wound closure at 12 weeks over SOC by 1.52 (credible interval: 1.37 – 2.22) in terms of relative risk (Treatment / SOC). This translates to an improvement of 8.53% (credible interval: 5.60%-19.7%) in percentage terms. In the ITT population, the IFSG +□SOC arm achieved a 47.6% closure rate versus 21.7% with SOC alone, a 25.9% absolute gain that was not statistically significant (n = 21, 95%□CI -0.02% to 0.493%, p□=□0.07, α = 0.05). In the ITT and PP population, IFSG +□SOC achieved a higher mean area reduction than SOC.

**Conclusion:** The interim analysis revealed that the IFSG products trended toward superiority over SOC. While the present interim analysis provides promising early results, limitations inherent to its preliminary nature warrant consideration. The alignment of these interim findings with the broader body of evidence reinforces biological plausibility and strengthens confidence that the final analysis will yield clinically meaningful results supported by high-quality evidence

## 1. Introduction

An estimated 4.5 billion individuals globally were affected by chronic venous disease stages C1-C6 in 2020, and approximately 0.1-0.3% of the world’s population developed a VLU [1]. Despite advances in treatment, approximately 7% of VLUs are estimated to remain unhealed after 12 months [2], and VLUs have a recurrence rate exceeding 70% after they are closed [3]. In 2022, the United States faced an estimated annual economic burden of over $4.9 billion for treating VLUs, covering costs related to healthcare practitioners, wound care products, inpatient hospitalization, medications, and compression therapy [4].

The use of biologically sourced matrices such as fish skin grafts represent a distinctive approach to the treatment of VLUs that leverages preserved extracellular components to support tissue regeneration. The intact fish skin graft (IFSG; MariGen®, Kerecis™ Limited, Isafjordur, Iceland) preserves naturally occurring components such as lipids, proteins, elastin, glycans, and other structural biomolecules. The graft is manufactured using a proprietary process and is indicated for the management of partial-and full-thickness wounds, including pressure ulcers, chronic vascular ulcers, diabetic ulcers, traumatic wounds, surgical wounds, and draining wounds.

The standard IFSG consists of a single-component graft created from the minimally processed skin of Atlantic cod (Gadus morhua) sustainably harvested from Arctic waters surrounding Iceland. The cod skin exhibits gross structural similarity to human dermis [5,6]. During processing, the skin is de-scaled, decellularized, lyophilized, packaged, and ter-minally sterilized with ethylene oxide, yielding a biocompatible, non–crosslinked, single-use medical device with a three-year shelf life.

## 2. Materials and Methods

THOR is a randomized controlled multicenter clinical trial designed to determine the difference in the proportion of subjects achieving complete closure of hard-to-heal VLU between Intact Fish Skin Graft plus standard of care (IFSG/SOC) versus SOC alone over 12 weeks. (clinicaltrials.gov #NCT06693570). This study was conducted at 7 SerenaGroup®, Inc. or affiliated centers throughout the United States with 136 patients with nonhealing VLUs. Enrollment for this study began October 2024 and interim analysis was conducted October 2025. The study population was drawn from patients suffering from VLUs who were attending wound clinics.

### 2.1. Objectives and Endpoints

The primary objective of the THOR clinical trial was to determine the difference in the proportion of subjects achieving complete closure of hard-to-heal VLUs between the IFSG/SOC arm versus the SOC alone arm over 12 weeks. The primary endpoint was the percentage of target ulcers achieving complete wound closure within 12 weeks.

An additional important endpoint evaluated was percentage wound area reduction from Treatment Visit 1 (TV-1) to Treatment Visit (TV-13) measured weekly with digital planimetry and physical examination.

### 2.2. Diagnosis

The diagnosis of VLUs relies on a detailed medical history, comprehensive physical examination, and selected diagnostic testing when indicated. VLUs most often occur in the gaiter area of the lower leg, particularly near the medial malleolus, and are commonly preceded by clinical evidence of chronic venous insufficiency such as edema, varicose veins, lipodermatosclerosis, and hemosiderin deposition [7]. These ulcers typically display irregular borders, a shallow wound bed with fibrinous exudate, and periwound skin changes including hyperpigmentation or stasis dermatitis. Pain intensity varies but is frequently described as a dull ache that worsens with leg dependency and is relieved by elevation.

A comprehensive clinical history is essential to distinguish VLUs from other chronic wound etiologies [8]. Key historical factors include previous venous disease, episodes of deep vein thrombosis, compliance to compression therapy, ulcer recurrence, occupational or lifestyle patterns involving prolonged standing, obesity, and prior interventions for wound care. The differential diagnosis should consider and rule out arterial ulcers, diabetic foot ulcers, pressure injuries, vasculitic lesions, and malignancy-related ulcers.

Bedside neurological evaluation was conducted to assess for loss of protective sensation. Vascular assessment was performed in all potential participants, with the ankle–brachial index (ABI) serving as the primary screening tool. Individuals with an ABI greater than 0.7 met inclusion criteria, whereas values above 1.3 prompted additional investigation for arterial calcification. In cases of incompressible or calcified arteries, commonly observed in patients with long-standing diabetes, alternative assessments such as the toe–brachial index (TBI; ≥ 0.6 indicating adequate perfusion) were used. A transcutaneous oxygen measurement (TCOM) of ≥ 40 mmHg was also accepted as evidence of sufficient perfusion..

### 2.3. Vulnerable Populations

Although vulnerable subjects were not specifically recruited for this study, vulnerable subjects were present in the potential subject pool.

### 2.4. Product Description

IFSG is a biologic tissue graft designed to facilitate tissue repair through preservation of the native extracellular architecture of fish skin. The graft retains a porous, three-dimensional collagen network that provides a scaffold for cellular infiltration, neovascularization, and granulation tissue formation.

The molecular components, including lipid, protein, and sugar components inherent to the material contribute to a physiologic healing environment, maintaining moisture balance and supporting host tissue integration. The flexibility of the graft allows it to conform closely to the wound bed, while its handling characteristics enable precise placement. IFSG is supplied as a sterile, single-use, sheet available in multiple sizes for use in a range of acute and chronic wound types.

### 2.5. Subject Characteristics

Individuals with nonhealing VLUs were recruited from participating wound care centers. After providing written informed consent in accordance with Institutional Review Board (IRB) approval and agreeing to comply with study procedures, participants underwent screening to assess eligibility. Inclusion and exclusion criteria used for enrollment are summarized in Table 1.

**Table 1.**
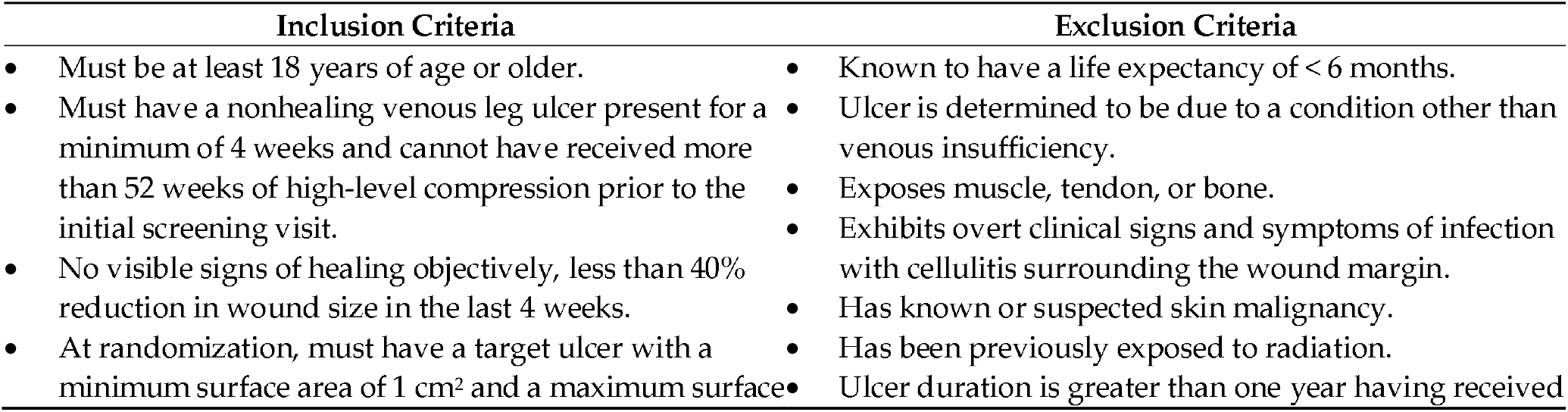
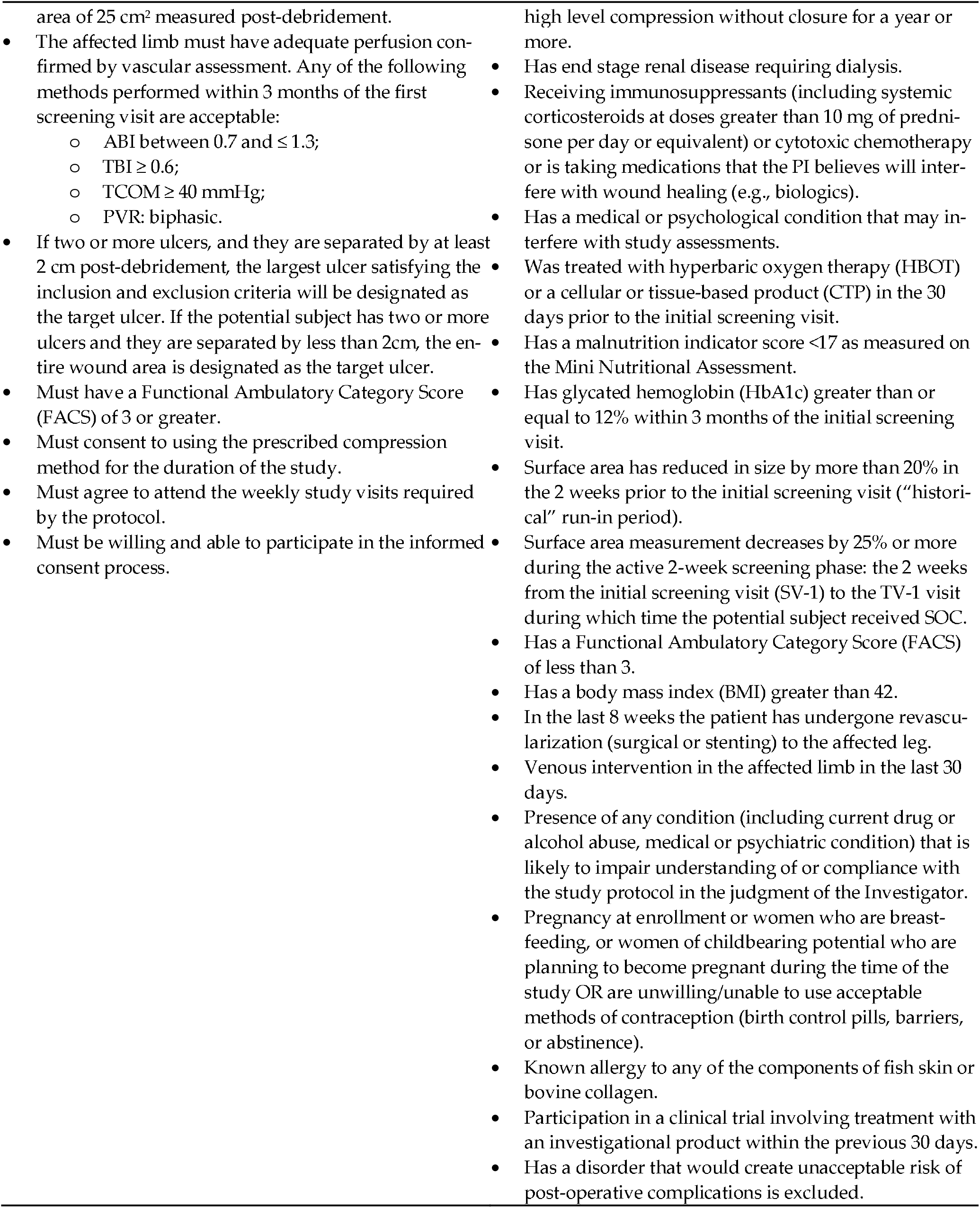
Inclusion and Exclusion Criteria.

**Table 2.** Study Schedule.

|  | SV | TV-1 | TV-2,<br>TV-3 | TV-4 | TV-5,<br>TV-6,<br>TV-7 | TV-8 | TV-9,<br>TV-10,<br>TV-11 | TV-12 | TV-13 | CCV |
| --- | --- | --- | --- | --- | --- | --- | --- | --- | --- | --- |
| Window Period | -14 | Day 0 | Week 1,<br>Week 2 | Week 3 | Week 4,<br>Week 5,<br>Week 6 | Week 7 | Week 8,<br>Week 9,<br>Week 10 | Week 11 | Week 12 | +14 |
| Record Medical History and Demographic Information | X |  |  |  |  |  |  |  |  |  |
| Assessment of Eligibility | X | X |  |  |  |  |  |  |  |  |
| Sign Informed Consent Form | X |  |  |  |  |  |  |  |  |  |
| Vascular Screening Test | X |  |  |  |  |  |  |  |  |  |
| Physical Exam | X | X |  |  |  |  |  |  |  |  |
| Mini Nutrition Assessment (MNA) | X |  |  |  |  |  |  |  |  |  |
| HbA1c | X |  |  |  |  |  |  |  |  |  |
| CEAP Classification | X |  |  |  |  |  |  |  |  |  |
| Fitzpatrick Scale | X |  |  |  |  |  |  |  |  |  |
| Historical Measurement | X |  |  |  |  |  |  |  |  |  |
| Randomization |  | X |  |  |  |  |  |  |  |  |
| Assessment for AE and SAE |  | X | X | X | X | X | X | X | X | X |
| Review medication for changes |  | X | X | X | X | X | X | X | X | X |
| Vital Signs | X | X | X | X | X | X | X | X |  |  |
| Wound Assessment | X | X | X | X | X | X | X | X | X | X |
| Pain Assessment (VAS) | X | X | X | X | X | X | X | X | X | X |
| wQOL |  | X |  | X |  | X |  | X | X |  |
| FWS |  | X |  | X |  | X |  | X | X |  |
| FACS |  | X |  |  |  |  |  |  | X |  |
| Study Ulcer Cleaning, Debridement (if applicable) | X | X | X | X | X | X | X | X | X |  |
| Study Ulcer Area with Imaging Device | X | X | X | X | X | X | X | X | X | X |
| Treatment Based On Randomization |  | X | X | X | X | X | X | X |  |  |
| Apply Dressing | X | X | X | X | X | X | X | X | X |  |

### 2.6. Study Procedures

Participants underwent a structured sequence of clinical visits including screening, treatment, healing confirmation, and follow-up phases to ensure accurate eligibility assessment, standardized wound care, consistent intervention delivery, and reliable endpoint determination. Subjects were evaluated weekly (± 3 days) over a 12-week treatment period, with any additional dressing changes recorded as unscheduled visits and abbreviated assessments performed when needed.

Participants who did not meet eligibility criteria at initial screening but were subsequently determined eligible were re-consented and assigned a new screening number. Up to three screening attempts were allowed, and those who subsequently met all inclusion and no exclusion criteria were enrolled. At the screening visit, conducted approximately 14 days prior to enrollment, informed consent was obtained, followed by a review of medical history to assess eligibility based on inclusion and exclusion criteria. Demographic data (including height, weight, BMI, gender, and ethnicity), medical and medication histories, and current use of non-steroidal anti-inflammatory drugs (NSAIDs) and opioids were recorded. A vascular screening test was performed unless recent results (≤3 months) were available. Vital signs were measured, and a general physical examination was conducted.

Additional assessments included the Mini Nutritional Assessment (MNA), HbA1c testing (unless available within 3 months), and condition-specific evaluations: Wagner grade, CEAP Classification, Fitzpatrick skin type, pain intensity via Visual Analog Scale (VAS), and detailed wound characterization (granulation tissue, nonviable tissue, depth, exudate, and periwound skin). To identify nonhealing VLUs, historical wound measurements from two weeks prior to screening were collected; a reduction in wound size of >20% during this historical run-in period resulted in screen failure.

During the two-week screening phase, standard-of-care (SOC) wound management included cleansing with normal sterile saline (NSS), sharp debridement, post-debridement ulcer photography and measurement using the provided imaging device, and application of silicone or foam dressings. Antiseptics were permitted during the screening phase but not during treatment. Skin protectants were used as needed for periwound maceration, and compression therapy was applied according to manufacturer guidelines.

At the enrollment/randomization visit (Treatment Visit 1; Day 0), eligibility was re-confirmed, medication changes were reviewed, and a symptom-directed physical examination performed. The percentage area reduction (PAR) over the screening phase was verified to remain <25%. Vital signs were taken, wound characteristics recorded, and questionnaires, including the VAS for pain, Forgotten Wound Score (FWS), Wound Quality of Life (wQOL), and Functional Ambulatory Category Scale (FACS), were completed. Eligible participants were randomized to receive IFSG + SOC or SOC alone. Wounds were cleansed, debrided, photographed, and measured using the imaging device prior to dressing application. Dressings were applied per protocol, with permission from the medical monitor required for any deviations or use of additional absorptive layers in highly exudative wounds.

Participants returned weekly (TV-2 to TV-12) for safety and efficacy monitoring, including adverse event assessment, medication review, vital sign collection, wound examination, VAS for pain, and administration of FWS and wQOL questionnaires at designated intervals (TV-4, TV-8, TV-12). SOC wound care or SOC wound care with IFSG treatment were performed at each visit, following identical cleansing, debridement, measurement, and dressing procedures.

At the final treatment visit (TV-13) or earlier if wound closure occurred, adverse events, medication updates, pain, FWS, wQOL, and FACS assessments were performed. For unhealed ulcers, wound characteristics were documented, and follow-up care arranged. Subjects with wound closure returned for a Closure Confirmation Visit 14 ± 3 days later, at which time adverse events, medications, pain, investigator confirmation of closure, ulcer imaging, and independent blinded verification were completed.

Subjects achieving closure were offered participation in a separate 12-month durability follow-up protocol, including in-person or telehealth visits at 3, 6, 9, and 12 months to confirm continued healing. Early withdrawals underwent final-visit procedures when feasible, and unscheduled visits were conducted for adverse event review, medication updates, and dressing changes as needed. At study exit, participants with unhealed wounds were transitioned back to physician-directed SOC. Independent confirmation of closure was performed by two blinded wound care specialists reviewing de-identified images from closure and confirmation visits, with discrepancies resolved in favor of the principal investigator’s assessment.

### 2.7. Subject Withdrawal

All participants had the right to withdraw from the study at any time during the treatment period without prejudice. The completion status of each participant’s involvement in the clinical trial was documented. In the event that study treatment or protocol-required observations were discontinued for any participant, the reason(s) for discontinuation were recorded. The investigator had the authority to withdraw a participant from the study at any time if deemed medically necessary. Whenever feasible, the reason for withdrawal or early termination was documented.

A participant was classified as lost to follow-up if they could not be reached after five telephone contact attempts and three written communications.

### 2.8. Subject Compensation

Participants received a reimbursement of USD 50 following completion of each study visit. This payment was intended to offset costs related to study participation, such as travel, parking, and the additional time required for visit procedures and data collection.

## 3. Results

A total of 136 VLU patients were screened from multiple sites and were evaluated in the interim analysis. Based upon Intent-to-Treat population, 34 patients received IFG plus standard of care, versus 34 patients in standard of care cohort. However, at the time of interim analysis, 10 patients from both treatment groups are considered ongoing and therefore cannot be evaluated. 3 patients were discontinued during the study and 43 were excluded during screening. This results in a total of 44 completed patients (21 IFSG+SOC and 23 SOC only). Summary statistics on demographic variables are provided in Table 3.

**Table 3.**
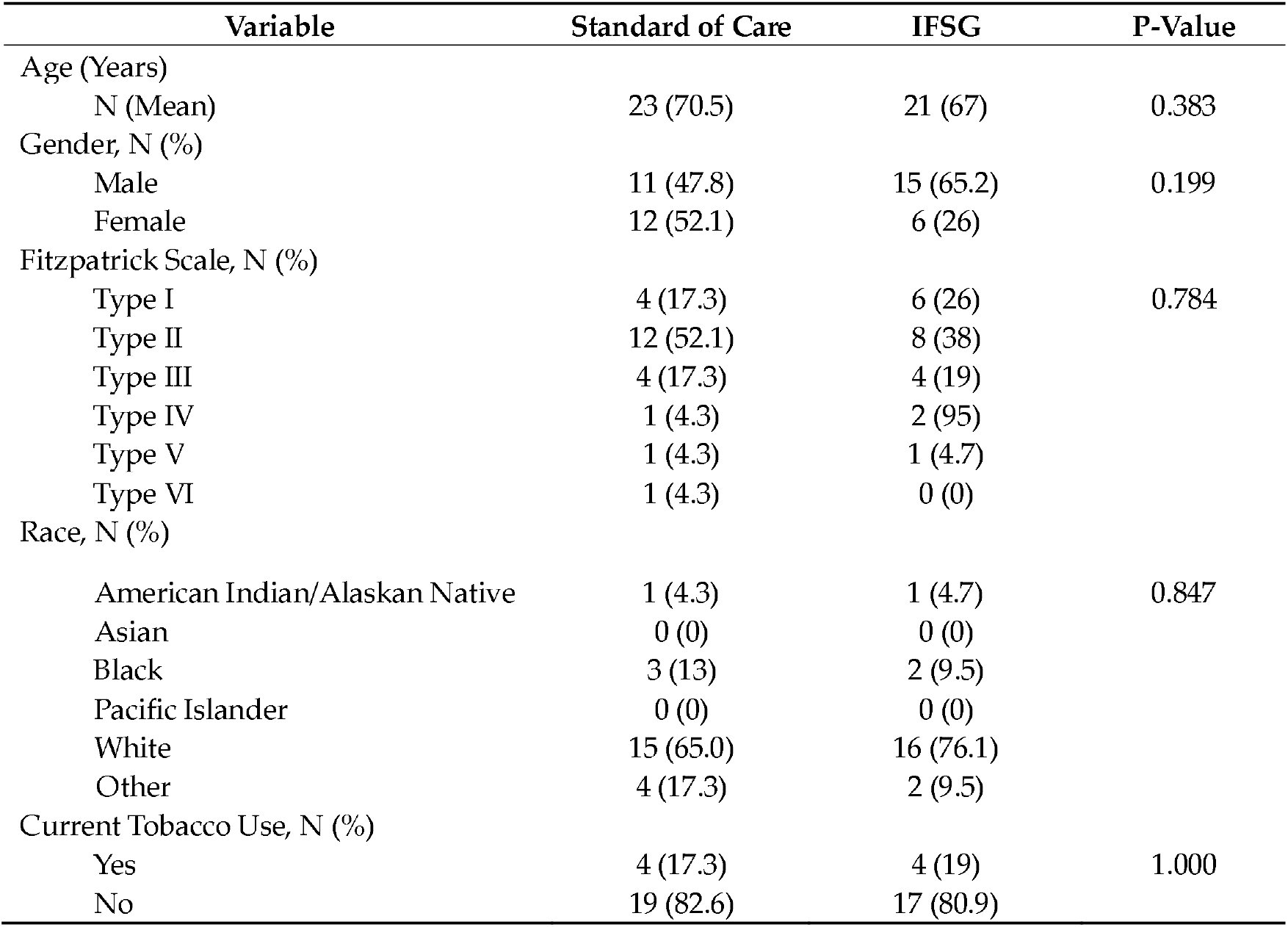
Demographic Summary Statistics by Treatment Group.

No statistically significant differences were observed across treatment groups (all p > 0.05), suggesting that randomization achieved adequate baseline balance. Wound area and wound age were used as stratification factors in the trial design, and at this interim analysis, they are summarized descriptively to assess balance, shown in Table 4. The reported p-values are exploratory checks of randomization balance and were not used to adjust the interim analysis endpoints.

**Table 4.** Stratification Summary Statistics.

| Variable | Standard of Care<br>(n = 23) | IFSG<br>(n = 21) | P-Value |
| --- | --- | --- | --- |
| Wound Area, N (%) |  |  |  |
| Less than 10 cm<sup>2</sup> | 20 (86) | 18 (85.7) | 0.488 |
| Greater than 10 cm<sup>2</sup> | 3 (13) | 3 (14.2) |  |
| Wound Age, N (%) |  |  |  |
| Wound > 60 Days | 5 (21.7) | 2 (9.5) | 1.000 |
| Wound < 60 Days | 18 (78.2) | 19 (90.4) |  |

The primary endpoint was assessed for the interim analysis. The primary endpoint is the percentage of target ulcers achieving complete wound closure in 12 weeks. Additionally, the percent area reduction (PAR) from TV-1 to TV-13 measured weekly with digital photographic planimetry, using an imaging device, and physical examination were analyzed. The intent-to-treat (ITT) and per protocol (PP) populations were analyzed.

In the ITT population, the IFSG +□SOC arm achieved a 47.6% closure rate versus 21.7% with SOC alone, a 25.9% absolute gain that was not statistically significant (n = 21, 95%□CI -0.02% to 0.493%, p□=□0.07, α = 0.05).

Among the PP population, closure achieved 44.4% on IFSG +□SOC versus 23.5% on SOC, a 20.9% difference, which likewise did not reach significance (n = 18, 95%□CI -0.1% to 46.8%, p□=□0.193, α = 0.05).

Within each arm, any individual PAR value falling below Q1 – 1.5*IQR or above Q3 + 1.5*IQR was flagged and excluded. For ITT, IFSG +□SOC outperformed Standard□of□Care on both average and median wound-area reduction, with a mean PAR ∼75% versus ∼41.6% (without outliers). Summary statistics are provided in Table 5.

**Table 5.** Percent Area Reduction (PAR) Summary Statistics without Outliers for ITT.

| Treatment Arm | n | Mean | Standard Deviation | Median | IQR |
| --- | --- | --- | --- | --- | --- |
| Standard of Care | 21 | 41.68 | 60.74 | 54.56 | 87.04 |
| IFSG + □SOC | 19 | 75 | 30.96 | 100 | 45.11 |

For PP, IFSG +□SOC outperformed Standard□of□Care on both average and median wound-area reduction, with summary statistics for each treatment group (without outliers) reported in Table 6.

**Table 6.** Percent Area Reduction (PAR) Summary Statistics without Outliers for PP.

| Treatment Arm | n | Mean | Standard Deviation | Median | IQR |
| --- | --- | --- | --- | --- | --- |
| Standard of Care | 16 | 51.51 | 65.04 | 75.05 | 63.84 |
| IFSG + □SOC | 17 | 74.66 | 31.88 | 100 | 46 |

Sequential images shown in Figure 1 document the trajectory of wound healing from SV-1, TV-1, and HCV in a patient assigned to the IFSG with SOC treatment arm.

**Figure 1.**
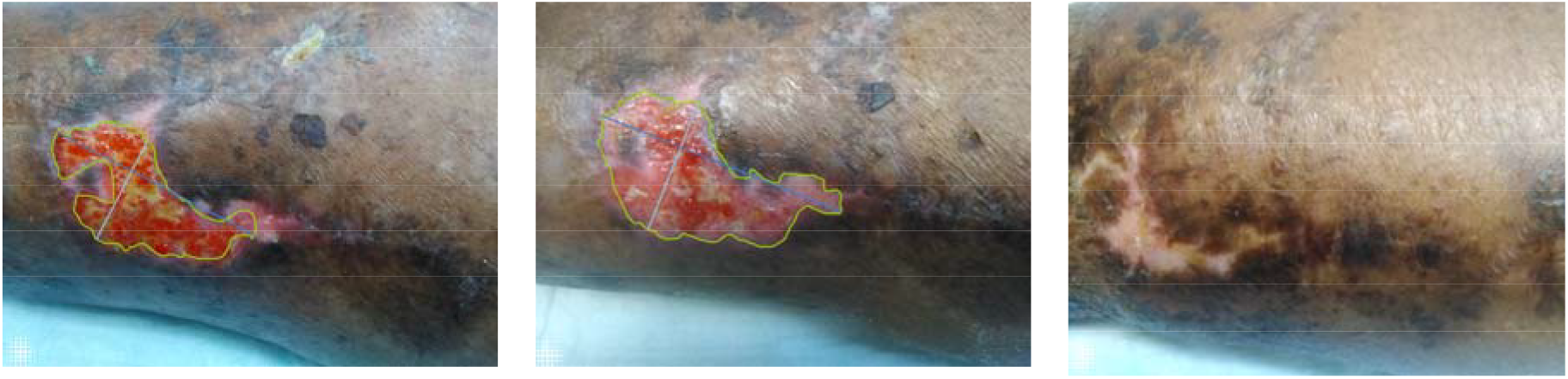
Digital images from SV-1, TV-1, and HCV (left to right), IFSG with SOC treatment arm.

## 4. Discussion

Interim analysis included a data lock on the electronic data capture (EDC) system and quality assurance review prior to data analysis. The purpose of this interim analysis is to determine balance across treatment groups and comparison to current standard of care for the primary endpoint and PAR. Patients were stratified by wound area, wound age, and patient age. There is no significant difference between strata between treatment groups, therefore, the randomization scheme achieved a balanced baseline. Additional analysis by the stratification group is planned for the final analysis.

For the primary endpoint, IFSG +□SOC was not statistically significant in the ITT or PP population. The small differences in sample size between populations may influence the results of the Chi-squared test, and additional enrollment will occur until the planned sample size is met for all treatment groups.

Percent area reduction provides insight into the closure rates by treatment group. In the ITT and PP population, IFSG +□SOC achieved a higher mean area reduction than SOC. This provides promising results at interim and confirmation of the clinical trial design prior to final analysis.

While the present interim analysis provides promising early results, limitations inherent to its preliminary nature warrant consideration. The current data lock represents fewer than half of the total planned enrollment (44 analyzed of a target 120 subjects), resulting in limited statistical power and wider confidence intervals than anticipated in the final design. Consequently, the precision of the effect estimates is constrained, and several endpoints did not achieve statistical significance despite clear directional trends favoring the IFSG arm. This pattern of an improved closure rate and greater percent area reduction suggests that the observed effect is unlikely to be spurious and may strengthen as additional participants complete the 12-week protocol.

Because this is an interim look, the certainty of the evidence under GRADE criteria is appropriately categorized as moderate, primarily due to sample size, incomplete follow-up, and interim reporting. However, the trial design of a prospective randomization, blinded endpoint adjudication, and multicenter enrollment mitigates major sources of bias and supports a trajectory toward high-certainty evidence once full accrual is reached. The direction of effect is consistent with previous published data on intact fish skin grafts in chronic and complex wounds, which have repeatedly demonstrated accelerated healing, improved granulation, and reduced treatment duration compared with standard of care. The alignment of these interim findings with the broader body of evidence reinforces biological plausibility and strengthens confidence that the final analysis will yield clinically meaningful results supported by high-quality evidence.

## Author Contributions

Conceptualization, T.S.; methodology, T.S., B.T., E.K. and Z.T.; data curation, D.S. and Z.T.; writing—original draft preparation, T.S., B.T., E.K., D.S. and Z.T.; writing—review and editing, T.S., B.T. E.K., D.S. and Z.T.; visualization, J.C.L.; project administration, B.B. All authors have read and agreed to the published version of the manuscript.

## Funding

The THOR trial was conducted under an unrestricted grant from Kerecis™ Limited

## Informed Consent Statement

Informed consent was obtained from all subjects involved in the study.

## Data Availability Statement

The data is proprietary but is available on request to the corresponding author.

## Declaration of Generative AI

During the preparation of this work the authors used OpenAI to support drafting and language refinement of portions of this manuscript, including improving clarity, grammar, and flow. The AI tool was not used for data generation, data analysis, interpretation of results, or drawing scientific conclusions. After using this tool/service, the authors reviewed and edited the content as needed and take full responsibility for the content of the publication.

## Notes

### Competing Interest Statement

John C. Lantis is the head of the Scientific Advisory Board for Kerecis, LLC and, as such, receives fair market compensation.

### Clinical Trial

NCT06693570

### Clinical Protocols

https://clinicaltrials.gov/study/NCT06693570

### Funding Statement

The study was funded by Kerecis LLC

### Author Declarations

Advarra Pro00081893

